# Expectations and attitudes in primary care towards home-based testing for diagnosing asthma: a mixed methods study

**DOI:** 10.1101/2025.11.14.25340253

**Authors:** Ran Wang, Katie Lawton, Binish Khatoon, Joanna Smith, Stephen J Fowler, Angela Simpson, Clare S Murray

## Abstract

**Background:** Asthma is frequently misdiagnosed because clinic-based tests miss its natural variability. As part of early stakeholder engagement, we examined primary-care healthcare professionals (HCP)’ views on using handheld spirometer and fractional exhaled nitric oxide (FeNO) devices for home-based diagnostic strategies.

**Methods:** This mixed-method study was conducted in two phases. Phase 1 data collection involved two focus groups with primary care HCPs using the Nominal Group Technique (NGT) to identify key priorities for home-based asthma diagnostic testing. These findings informed the development of a national electronic survey distributed to primary care HCPs during Phase 2.

**Results:** Using the NGT focus group participants (n=21), we identified key advantages, challenges, and facilitators for implementing home-based asthma diagnostics, which informed the development of an e-survey. Of 235 HCPs who consented to start the survey, 104 completed all 10 questions. Respondents represented a wide demographic and practices across all levels of socioeconomic deprivation. Only 3% considered home-based diagnostics strategy is unlikely to be feasible. The most frequently cited barrier was high device cost, while patient engagement and device accessibility were identified as the most important enablers. Most respondents highlighted faster and more accurate asthma diagnosis as key potential benefits.

**Conclusion:** Home-based asthma diagnosis using handheld spirometry and FeNO is generally viewed favourably by primary care professionals based on survey findings, though implementation challenges are multifaceted. Success will require system-level changes in how home-based testing is delivered and supported. The subsequent phase involves evaluation of test feasibility and accuracy, followed by assessment of clinical and cost-effectiveness.

## INTRODUCTION

Asthma is a chronic disorder of the airways, affecting 10% of the UK population (NHS Digital, 2019). It is characterised by reversible airflow obstruction and airway inflammation, with patients typically experiencing one or more symptoms such as wheeze, breathlessness, chest tightness or cough.

Asthma misdiagnosis occurs in a third of patients labelled with the condition (Aaron et al., 2017; van Schayck et al., 2000). The hallmark of asthma is the temporal variability in its underlying pathophysiology, including fluctuations in airflow obstruction and airway inflammation (Wang et al., 2021b); over 74% of patients experiencing worsening symptoms overnight (Turner-Warwick et al., 1988). It is therefore unsurprising that the current one-off, clinic-based testing during the day is ill-suited to capture this inherent variability. Indeed, there is now mounting evidence underscores the significance of the timing of test performance, such as spirometry bronchodilator reversibility tests and fractional exhaled nitric oxide (FeNO, a biomarker of airway inflammation), in influencing diagnostic outcomes (GINA 2025, Wang et al., 2024, Wang et al., 2021a, Wang et al., 2023, Knox-Brown et al., 2025). Specifically, performing FeNO and spirometry, the two first-line asthma diagnostic tests recommended by the joint BTS/NICE/SIGN 2024 asthma guidance (NICE 2024) in the morning lead to a higher likelihood of positive results compared to tests performed in the afternoon (Wang et al., 2024, Wang et al., 2021a, Wang et al., 2023, Knox-Brown et al., 2025). Notably, the Global Initiative for Asthma (GINA) 2025 strategic report also recommends performing diagnostic testing when patients are symptomatic. Given constraints on primary care resources, this is unlikely to be widely achievable. Therefore, improving asthma diagnosis will require innovative approaches.

In the current routine primary care, the only method that incorporates variation is home-based peak expiratory flow (PEF) diurnal monitoring, a test introduced more than 60 years ago, which has been in favoured for its simplicity and low cost. However, in contrast to forced expiratory volume within one second (FEV_1_) measured by spirometry, PEF is an insensitive measure of small airway obstruction (Goldberg et al., 2001) and therefore offers limited diagnostic utility (with a sensitivity of 15%, Simpson et al., 2024). With the advancement in technology, remote spirometry and FeNO testing have become possible using hand-held devices.

In the context of the UK Governments’ 10-Year Health plan to shift from analogue to digital care (DHSC., 2025) and initiatives such as NICE’s Early Value HealthTech programme for respiratory diagnostics (NICE., 2025), digital technologies have been shown to improve asthma control and quality of life (Khusial et al., 2020) and facilitate asthma monitoring (Wang et al., 2023). However, their role in enhancing asthma diagnostic accuracy remains unknown. As most asthma diagnoses are made within primary care, assessing the acceptability of this testing approach and identifying potential enablers in this setting is the first step to evaluate its potential clinical utility.

Our objective was to understand early-stage stakeholder perspectives on a home-based diagnostic approach for asthma. We specifically examined primary-care healthcare professionals (HCPs)’ expectations, motivations, barriers and key enablers to adopting home spirometry and FeNO in the asthma diagnostic processes.

## METHODS

### Study design

This is a mixed method study conducted in two phases. Phase 1 comprised of focus group activities with primary care HCPs. Using the Nominal Group Technique (NGT) (Delbecq and Van de Ven, 1971), we collected information that informed the development of a national e-survey; survey responses were collected during phase 2.

This study was linked with the Rapid Asthma Diagnostic Clinics for Asthma study (RADicA, https://www.radica.org.uk) (Murray et al 2024) and conducted in parallel with a feasibility study evaluating home spirometry and FeNO for asthma diagnosis. Patient acceptability is reported elsewhere (Khatoon et al., 2025).

### Phase 1: Focus group activities

Primary-care physicians, advanced nurse practitioners and community nurses from local primary-care networks who are involved in the diagnosis or care of asthma patients were recruited through local primary care networks (PCN). Health professionals without experience in providing asthma care or not working in primary care settings were excluded.

We purposefully sampled participants from a diverse background, working at different geographical locations in Greater Manchester with varied socioeconomic status, and different asthma diagnosis and management experiences. Participants were recruited via two sources: 1) snowball sampling, whereby initial participants referred additional participants until group size was saturated, to maximise geographical spread (Focus Group 1, [FG1]) and 2) advertising across five local PCN practices to include a wider range of healthcare professional roles, albeit with narrower geographical coverage (Focus Group 2, [FG2]).

Both focus groups were undertaken in person between Oct 2023 and Feb 2024. Participants were allocated to groups according to their geographical locations, with 9 and 12 in each group respectively. The NGT was used to structure the focus groups, and is a well-recognised structured group decision-making process, which supports small groups of participants to generate and prioritise ideas in response to a question with the ultimate aim of gaining group consensus (Delbecq et al., 1975; Harvey & Holmes, 2012). We used the NGT to generate and prioritise questions for a national survey, designed to understand the perceived advantages, barriers, enablers for using home diagnostic devices for diagnosing asthma. To minimise bias, focus groups were led and facilitated by BK (an experienced qualitative researcher without medical background) and KL (GP with qualitative research experience), respectively, in the absence of the clinical study team. The format of the focus groups was in keeping with NGT methods: *Silent idea generation, round robin sharing, discussion and clarification, ranking and consensus*. Detailed format of group sessions is described in the online supplementary material (Section E1). Group sessions lasted approximately three hours. The group activities were also audio-recorded and transcribed, with permission of all the participants. Example quotes presented in the study reflect both written responses from the silent idea generation phase and verbal contributions during the sharing, discussion and clarification stages.

### Phase 2: National survey

The key potential barriers and enablers, advantages and disadvantages developed from the focus groups informed the construction of an online survey. The e-survey was circulated via primary care WhatsApp and Facebook groups, email distribution through primary care networks and GP trainee groups and the Primary Care Respiratory Society’s InTouch newsletter. Responses were collected between June 2024 and August 2024.

This study was approved by the University of Manchester Research Ethics Committee (HEARER Study, 2023-17916-31137). All focus group participants provided informed written consent. Informed consent to survey completion was implied by participants’ decision to complete the survey and answered “yes” to the first survey question “Do you agree to take part in completing the survey?” (Online supplement, Table E1).

### Data analysis

Data were collected and analysed during the NGT process (Section E1). For each domain (advantages, disadvantages, barriers and enablers), participants individually ranked the top ten prioritised items based on their perceived importance before submitting their responses. The aggregated rankings were then calculated and shared with the group. The top ten ranked items for each domain were then selected for inclusion in the survey; where rankings were tied, all tied items were retained, resulting in some survey domains containing more than ten items.

To report more comprehensive findings from the focus group discussions, content analysis (Elo & Kyngäs, 2008; Braun & Clarke, 2022) was undertaken with the focus group audio data. The process included:

1. Transcription of the audio recordings by KL
2. Familiarisation of the data through repeated reading of the transcripts and group, and re-listening to the audio recordings notes by BK and KL
3. Closely related or overlapping responses were synthesised into overarching themes through an iterative process. Duplicate items were removed to avoid redundancy.

### Survey analysis

Descriptive statistics were used to summarise survey response items. Survey responses from participants who completed all questions were included in the primary analysis. As a sensitivity analysis, demographic characteristics and rankings of the importance of advantages, disadvantages, barriers and enablers were also analysed using data from all individuals who responded to each respective question, regardless of survey completion. Missing data were excluded. All statistical analysis were performed using R Version 4.2.2 (Rstudio 2022.12.0). Responses to the free-text questions in the survey were analysed using content analysis by KL (details are included in Online supplement, Section E2).

## RESULTS

Primary care HCPs (Table 1) working across geographical locations with a mix of urban, sub-urban and rural area in Northwest England were recruited. The catchment areas of their clinical practices covered some of the most deprived areas within the UK.

**Table 1.** Primary care HCPs demographics.

| Focus groups | Professional roles | Number |
| --- | --- | --- |
| FG1 | GPs | 8 |
|  | Practice nurse | 1 |
| FG2 | Healthcare assistants | 2 |
|  | Practice nurse | 2 |
|  | GP trainees | 2 |
|  | GPs | 6 |

While top ten rankings were highlighted, we incorporated these into the two broad themes, giving a more holistic picture of the overall pattern of responses.

### THEME 1: The potential benefits of using home diagnostic strategies

The first of the two themes reflected professionals’ views about the potential benefits of home testing devices or motivators, including the *advantages* and *enablers* of home asthma testing.

The key advantage of home asthma testing was the potential to enhance accuracy of asthma diagnosis. Health professionals perceived that improved diagnostic accuracy would optimise the use of health resources - saving time and money and reducing unnecessary referrals to specialist services. Furthermore, home testing could increase the number of patients with asthma receiving appropriate treatment, improve health outcomes, and reduce inappropriate prescribing in those misdiagnosed. Home testing could become resource-sparing for both primary and secondary care.

> *‘Get the right diagnosis (or lack of one) faster; diagnostic certainty should save money and time by avoiding unnecessary treatments’. FG2*
>
> *‘Reduces unnecessary prescribing; reduces steroid need’. FG1*
>
> *‘I do wonder whether we push people up and up and up through the different levels on inhalers when the diagnosis isn’t actually secure, so if we can actually confirm asthma or not out of these tests then we improve outcomes, symptoms and costs’. FG2*
>
> *‘Less use of resources, for example clinic rooms, and done in patient’s own time rather than a nurse appointment*.*’ FG2*

Furthermore, a home testing strategy could enhance professionals’ confidence in an asthma diagnosis. For example:

> *“The knowledge that you are providing better patient care”. FG1*
>
> *“It will make us feel more confident in making a diagnosis or deciding they do not have asthma”*. FG2
>
> *‘Clinicians more reassured of the right diagnosis” “Patients should be more confident in the diagnosis’. FG2*

Health professionals postulated that home testing, compared to current practice, could support greater patient empowerment and offer a more patient-centred approach to asthma care. The value of home testing across all ages was highlighted, for example:

> *‘Patients are more engaged/invested in their own care’. FG2*
>
> *‘Might help to diagnose young children at home, they might be more compliant at home than in the surgery’. FG1*
>
> *“Patients who are more engaged in their own conditions, who are more empowered”. FG2 “Those who feel they are more part of the process are more likely to invest”*.FG2
>
> *“Better patient understanding of their own disease. This may enable them to get more involved”*. FG2

The most frequently discussed enabler of home asthma testing was the availability of a training package for staff, which reflected both knowledge and confidence in interpreting the test results. Similarly, professionals highlighted that patients would also need additional materials/support to assist in completing the tests accurately, especially those identified as likely to struggle. Examples included:

> *“Training for staff, for example on how to interpret the results, otherwise the GPs just won’t refer people to have the test done”*. FG1
>
> *“Providing clear instructions and written/translated literature such as leaflets, texts and videos”*. FG2
>
> *“Ensuring patients understand the test and the benefits there could be in symptom reduction”*. FG2

Across both focus groups, some HCPs felt that clear pathways and supporting infrastructure covering device issuing and training, results interpretation, treatment decisions, and administrative support could streamline the service. For example:

> *“A clear pathway for issuing devices, returning devices, interpreting results and then discussing results with the patient”*. FG2
>
> *“I suppose it depends how each practice ran it, some might send a video with a link on how to use the equipment which is no extra time, or some might want to invite patients in to demonstrate to them which would take more time”*. FG2
>
> *“The ability to import the results directly into the patient records could save time”*. FG1
>
> *“An algorithm or a report for the results which then told me what to do next i*.*e. what inhalers to use. Yes, a service which gives the results with the conclusions, like the remote ECG service some practices use”* FG1
>
> *“A pharmacist to prescribe according to the results”, “A central service that tagged the machines, called patients about it and did the admin side of it”*. FG1

Other HCPs perceived that a key enabler to home testing would be for secondary care services to deliver the diagnostic tests. For example:

> *“A hub to refer into, rather than us doing it in primary care. Lots of PCNs are doing that, having a centralised hub for example for respiratory testing”*. FG1

Professionals highlighted that providing financial support for healthcare organisations to purchase devices would enable widespread use in practices. Financial solutions included devices free of charge to NHS services including replacement costs of broken/lost devices, or incentivised by Quality and Outcome Framework [QOF]. Examples include:

*“QOF recognition, i*.*e. financial remuneration”*. FG1

*“The ability to offer an incentive to those who return a device”. FG2*

*“Cheap smart phone for those who do not have one to loan out”*. FG1

*“Having enough devices available to provide to public”*. FG2

### THEME 2: The challenges of using home diagnostic strategies for asthma

The second of the two themes reflected professionals’ views about the potential challenges of home testing, derived from the discussions of *the disadvantages and barriers*.

The most frequently highlighted disadvantage for home testing was that the devices could be a strain on resources available in primary care settings. This was a particular concern when HCPs discussed the current climate of rising financial burden within healthcare. For example:

> *“Cost of the equipment and malfunctions. High start-up costs”*. FG1
>
> *“Cost to the practice if not returned or broken”*. FG2
>
> *“Who would fund the devices? The GP practice or PCN? Our practice gets paid for doing spirometry so would doing this cause a potential loss of income. Our practice is the only one in the whole PCN that does spirometry”*. FG1

A further disadvantage discussed across both focus groups was the perceived amount of time and commitment teams would need to give to developing this service, which may divert away from other services. Examples include:

> *“Increased clinician time to go with it, which covers everything from showing the patients how to do it and then the nurse or doctor or whoever looking at the results. Also, someone is going to have to inspect the devices, quality check and clean the devices in-between clients, which is more time. This all comes under increasing burdens on primary care really”*. FG2
>
> *“Organisational/admin burden”*. FG1
>
> *“Longer appointments might be needed, meaning that other patients miss out on other services”*. FG1

A number of HCPs emphasised that many primary care services may not be ready or have the capacity to incorporate new technology into their practice. Furthermore, there was some scepticism around how much home-based diagnostic strategy would improve the current practice. Examples quotes included:

> *“Diagnosis may still be no faster than the current situation, so what does this actually change” Focus group 2*
>
> *“If there is high demand, would there be enough machines to reduce waiting times anyway, so would there be any benefit to the new system, or would it be just as quick to diagnose them the way we are already. We have one FeNO machine in our practice which has an 8-week waitlist, so we might as well diagnose them the old way”*. FG1

HCPs, particularly those working in more deprived areas, were concerned that the devices may not be as accessible to certain patient groups, for example those whose first language is not English, the elderly, those with cognitive impairment or those without smart phones.-potentially exacerbating health inequalities. Example included:

> *“Accessibility could be an issue, with patient understanding and also access to smart phones, we have a lot of elderly patients who don’t have smart phones so we would still need to book spirometry”*. FG1
>
> *“I think a lot of illiterate people would struggle with this, a lot of our patients cannot read and write”*. FG1
>
> *“If it is not well implemented it could increase health inequalities by being more available in richer or whiter areas”*. FG2

HCPs shared concerns related to the confidence of both patients and clinical staff to use and interpret the devices. In addition, there were concerns about increased burden to patients. Example quotes include

> *“Patient time, patient compliance and remembering to do the tests”* FG2
>
> *“Needs reliable, good technique from patients which I am not sure they have”*. FG1
>
> *“One problem is relying on patients to record the readings”*. FG2
>
> *“There is a lack of clinician expertise to interpret FeNO and spirometry results”*. FG1

### Phase 2: Primary care health care professionals national e-survey

A national e-survey was developed from the Phase 1 NGT activities (Table E1). Of the 235 primary care HCPs who started the e-survey, 104 completed all questions. Respondents reflected diversity in professional role, geographic distribution and area-level deprivation and variation in experiences in asthma diagnosis (Figures 1-2, Figure E1). Demographics were similar between those who started but did not finish the survey and those who completed it (Figure 1-2 and E2, Table E2). Over half of primary care healthcare professionals reported reviewing patients presenting with asthma-like symptoms on a weekly basis; 92% indicated they do so at least monthly. Over 96% of HCP survey respondents stated home diagnostic testing in primary care may be implementable (Figure 3).

**Figure 1.**
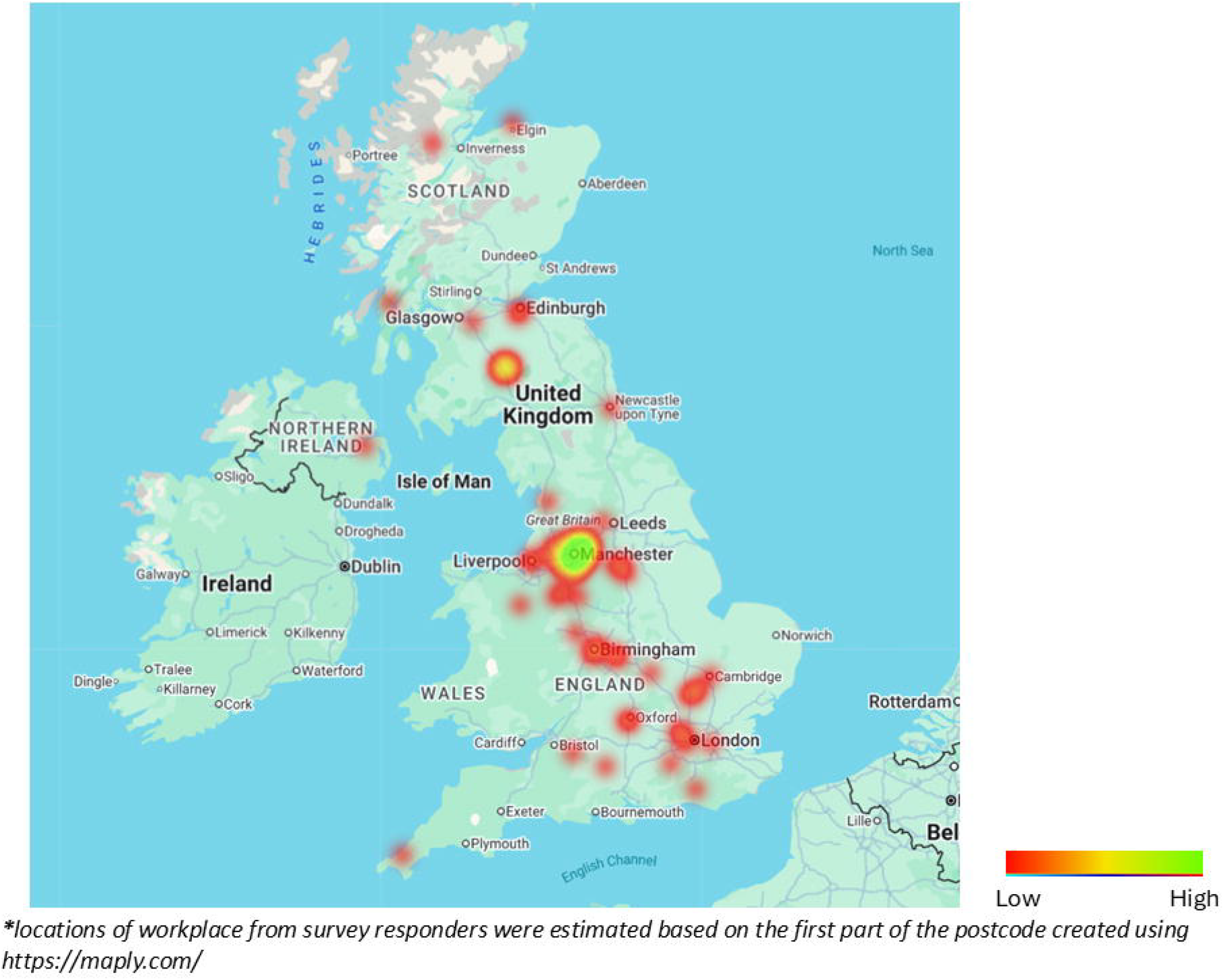
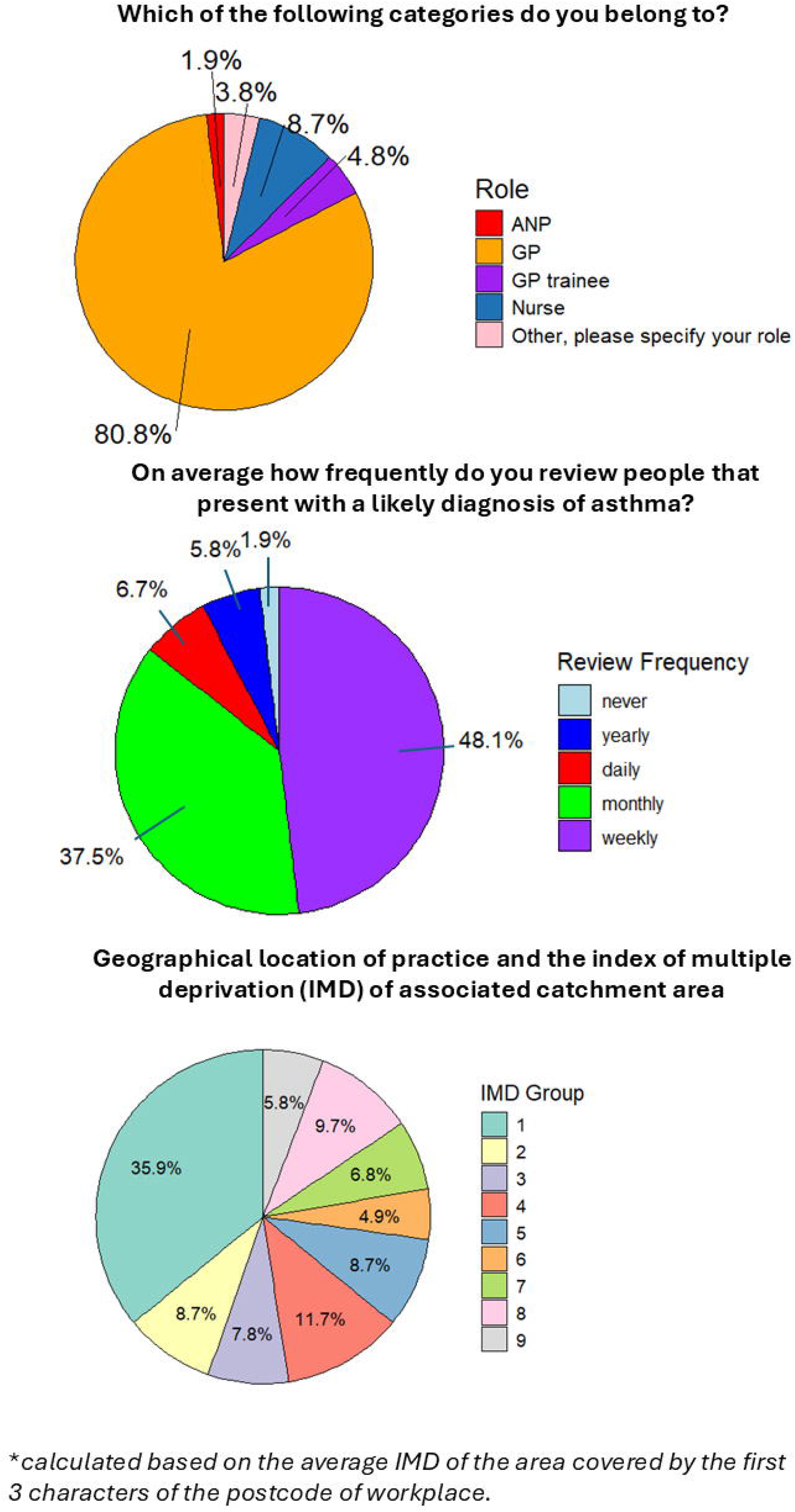
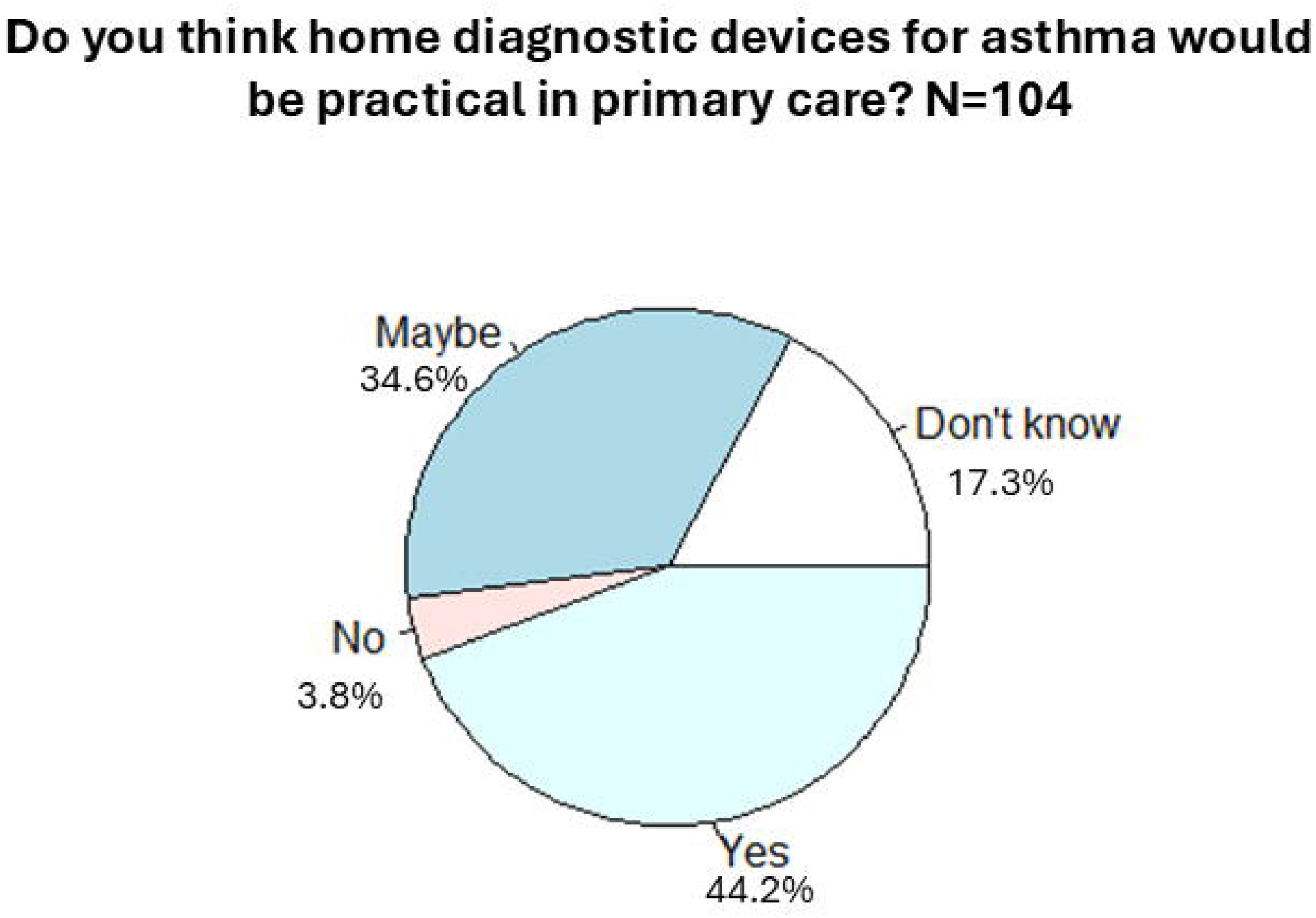
Geographical locations of survey responders (n=104)^*^.

**Figure 2.**
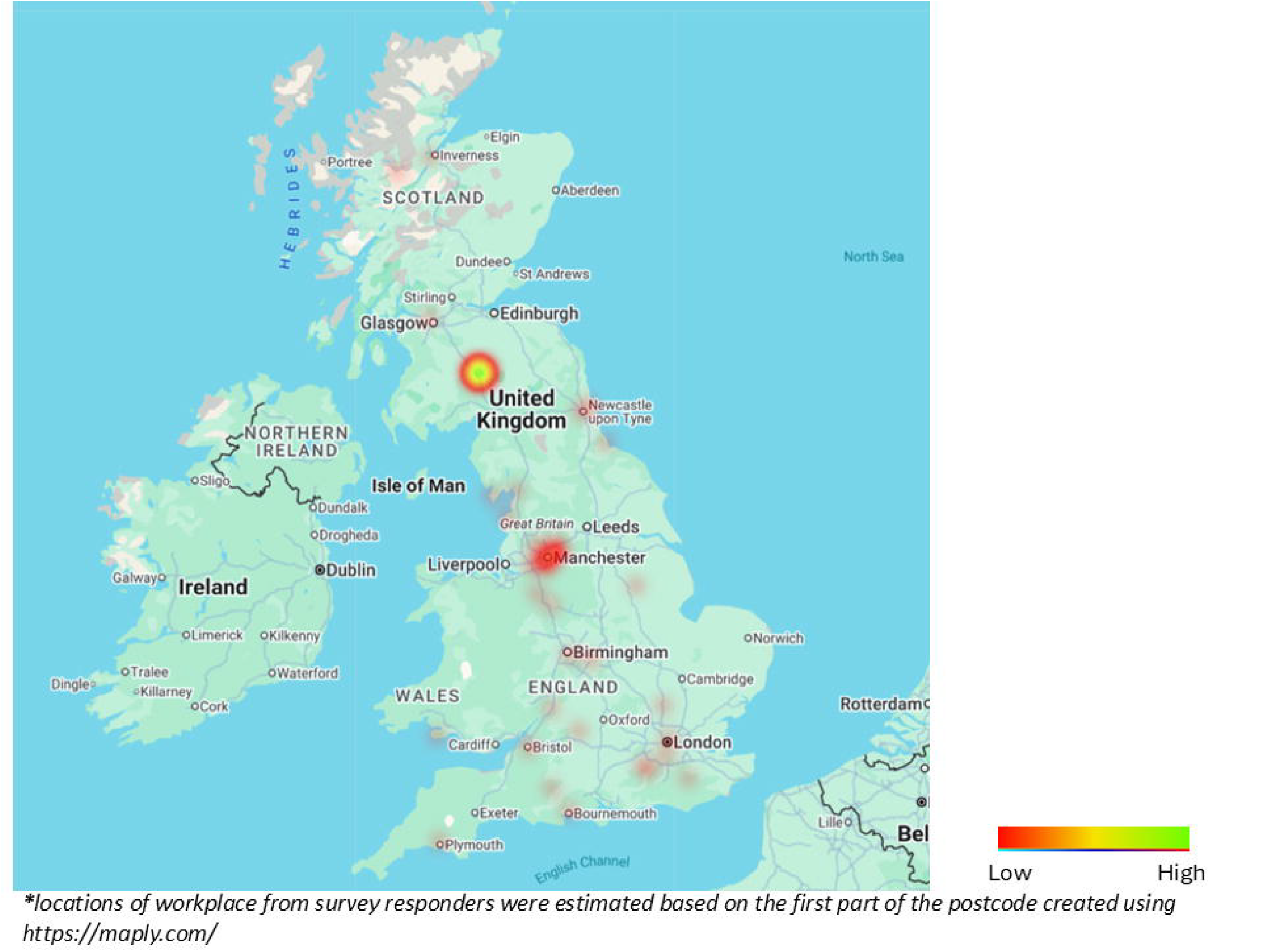
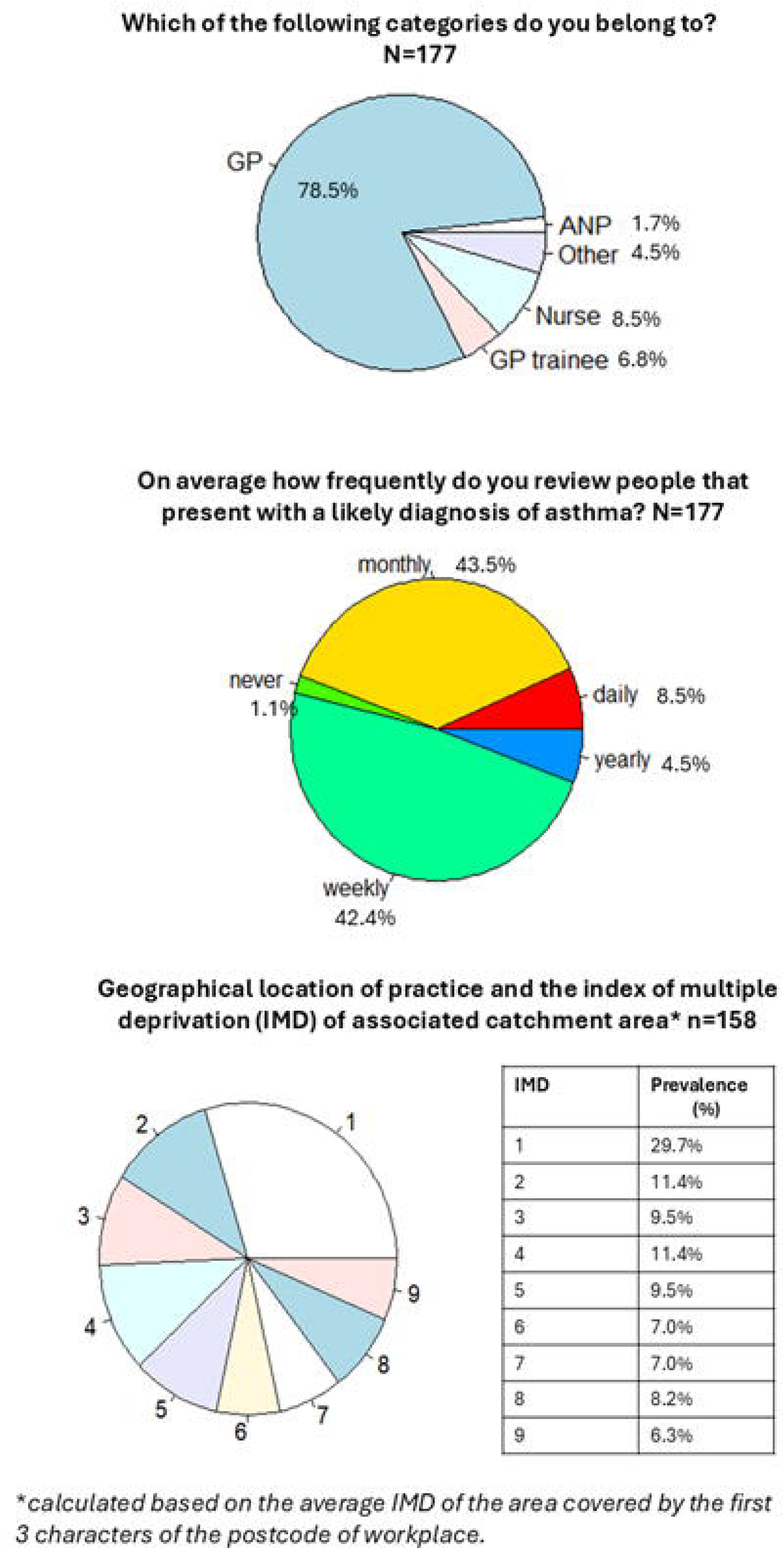
Survey respondents’ demographics of those who completed all survey questions (n=104).

**Figure 3.**
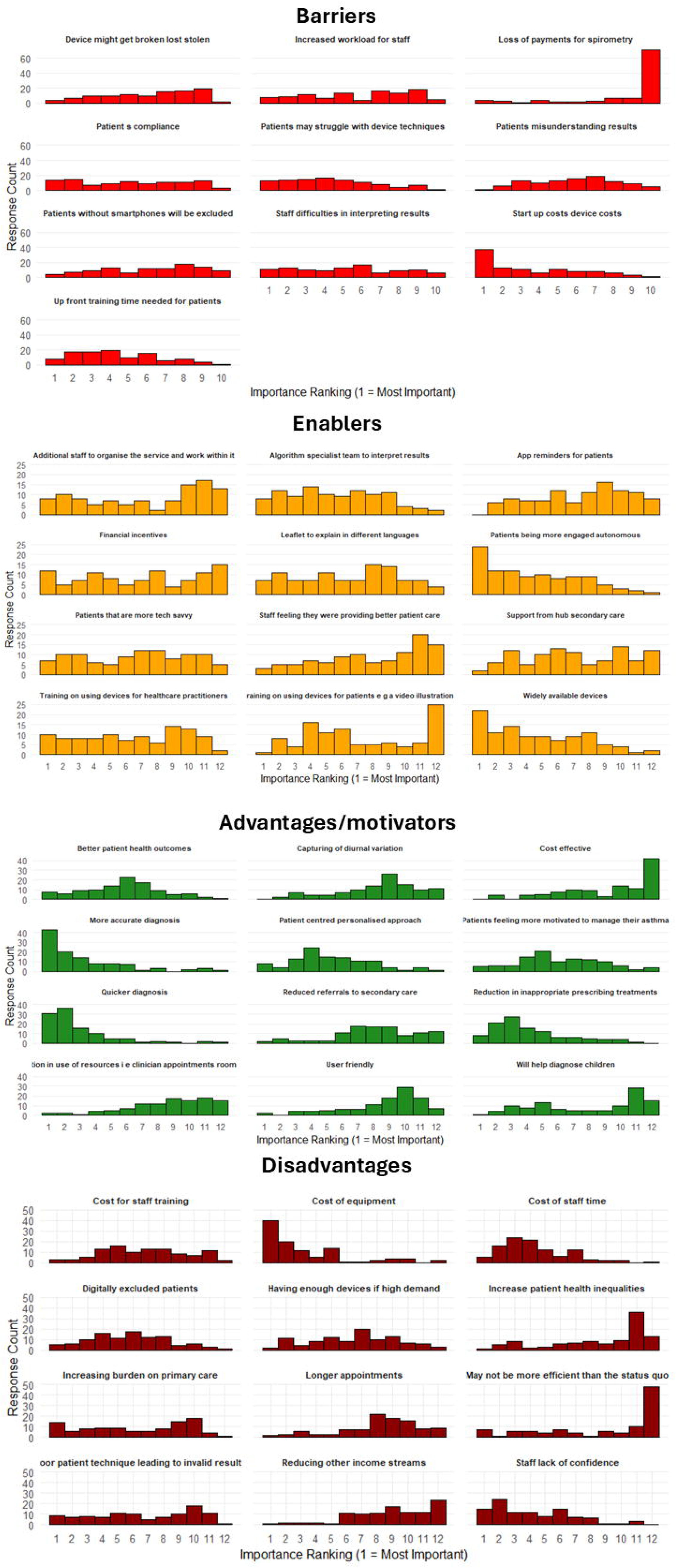
Feasibility for home-based testing in primary care: “Do you think home diagnostic devices for asthma would be practical in primary care?” (n=104)

The responses were heterogeneous, and the importance placed on different factors varied across healthcare professionals. Whilst high device costs were most commonly ranked as the primary barrier, patient engagement and the availability of widely accessible devices were viewed as key enablers to implementation (Figure 4, Figure E3). Improved accuracy and faster diagnosis of asthma were most frequently rated by HCPs as an important potential advantages of home testing.

**Figure 4.**
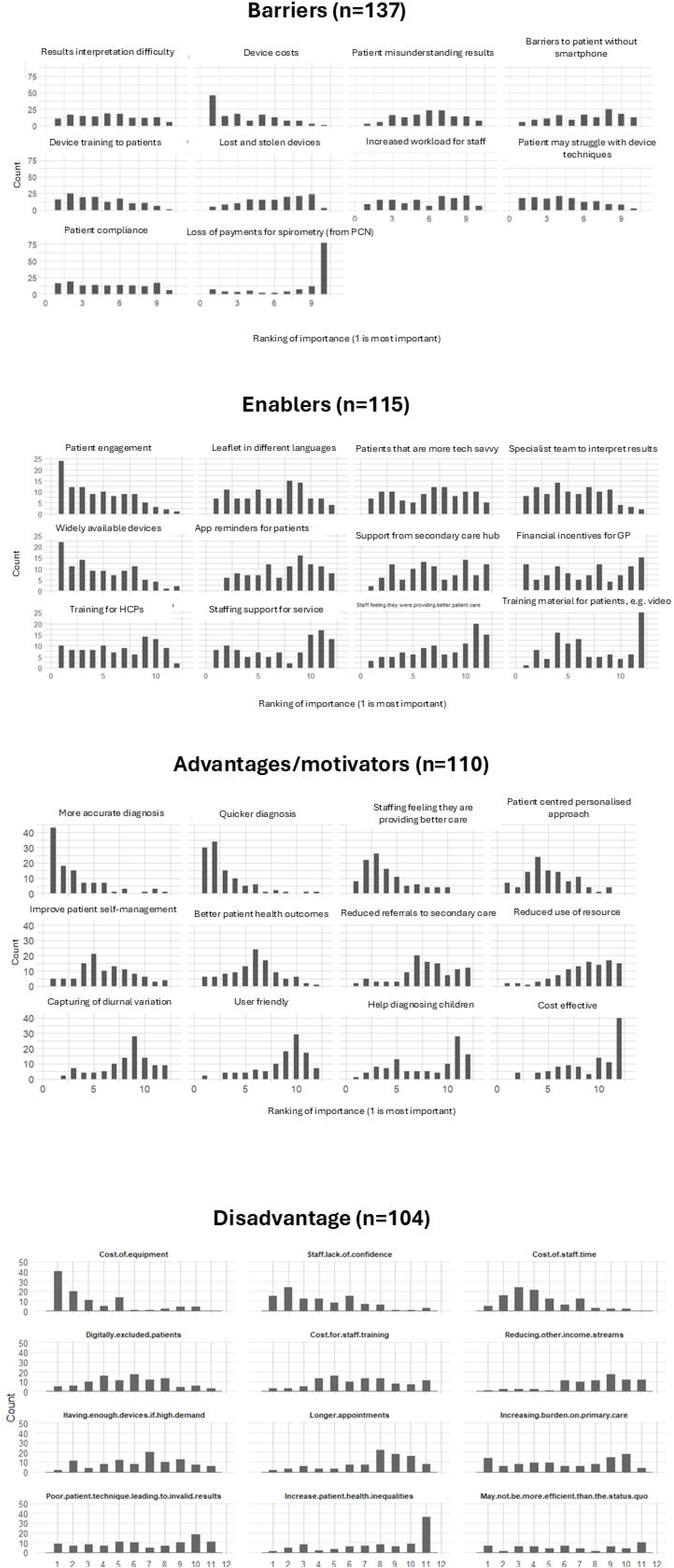
The ranking of the importance of potential advantages (motivators), barriers and enablers reported by e-survey (n=104); 1 is the most important.

Forty-four percent (n=104) of survey participants responded to the open text question: “In your individual practice what would be the most important factor that would enable/help you to use home diagnostic devices for asthma?”. The responses were grouped into three priority areas: resources, training and support. Responses relating to having enough resources to implement home-testing were most common (n=62 votes), ranging from funding to provide staff time and appointments to deliver the service, device related costs and accessibility. The second most common priority area focused on HCP training (n=28), including use of the devices and results interpretation. The third priority area related to support in rolling out the new service (n=15); ranged from support from local Integrated care boards (which included financial incentives, or extra staff to help with the process), to device support for patients and support from local respiratory teams with the interpretation of results.

## DISCUSSION

The potential utilisation of digital devices for home-based asthma diagnostic testing was generally well received by healthcare professionals in primary care based on survey results. However, the successful implementation of such technologies is challenged by a range of potential barriers. Key enablers, including adequate training, equitable access to devices, and sustained patient engagement, are critical to ensuring the efficacy and implementability of this clinical approach.

Our findings were consistent with previous studies: Miles et al (2017) demonstrated that although digital technologies are embraced by patients, carers and healthcare professionals for the management of asthma, sufficient training, education and support must be in place to ensure the feasibility and efficiency of this strategy. Van de Hei et al. (2023) conducted a study exploring the multi-stakeholder (patients and healthcare professionals) capacity and needs of smart inhaler use for improving asthma adherence. They found that enhanced asthma care and cost savings were contingent upon the technology being user-friendly and accompanied by adequate training and education for both patients and staff. Key barriers identified included the lack of reimbursement for additional workload and concerns regarding the security of data storage. Interestingly, the barriers to objective testing in airways diseases in primary care are complex (Yamada et al., 2022) even for established methods such as laboratory-based spirometry. These barriers include similar domains, such as the lack of skills and knowledge in test performance and result interpretation and limited test accessibility; test appointment non-attendance (lack of patient engagement) was also highlighted (Yamada et al., 2022).

Inequalities in access to asthma diagnostics remain a significant barrier to timely and accurate diagnosis, particularly among socioeconomically disadvantaged and minority populations (Forno and Celedon, 2009, Akinbami et al., 2017). Language barriers, digital literacy, and healthcare infrastructure gaps may further compound these disparities, contributing to delayed or missed diagnoses and suboptimal disease management (Flores, 2006). Emerging home-based digital health technologies, including handheld diagnostic tools, have the potential to reduce some of these barriers by decentralising testing. However, without careful implementation that accounts for affordability, digital access, and cultural and linguistic appropriateness, such innovations risk exacerbating rather than alleviating existing inequalities (Veinot et al., 2018). Ensuring equitable asthma diagnostics will require targeted strategies to engage underserved populations, subsidise device provision and deliver training and support that is inclusive and accessible to all.

### Limitations

Although the focus group discussions involved a broad sample of primary care healthcare professionals, these were limited to two sessions, making it unclear whether data saturation was achieved. The composition and diversity of professional roles of each group differ, likely due to different recruitment strategies. Snowball sampling captured a wider geography with less role diversity, whereas local advertisement through PCN captured greater role diversity within a limited geography. The two recruitment strategies therefore provide complementary strengths. However, we acknowledge that this imbalance, particularly in FG1, may have shaped discussion dynamics and constrained the depth and breadth of perspectives from minority roles. Although the facilitator used structured turn-taking and targeted prompts to mitigate dominance, residual risk of under-representation remains. Nevertheless, the national e-survey captured responses from a wider and more diverse population, with no further information emerging from the open-ended responses, suggesting good thematic coverage. The e-survey’s response rate cannot be reliably estimated because the number of HCP reached was unknown, and completion was modest. Variable familiarity with home-based spirometry/FeNO devices and the concept of home-based testing may have affected survey responses. It is also important to note that HCPs who participated in the focus groups or completed the e-survey may have had a greater engagement in asthma care, potentially leading to selection bias. Furthermore, we observed a high survey dropout rate, potentially introducing further bias. However, we found no difference in demographic data between survey respondents who completed the survey and those who did not. Although focus group participants received demonstrations of the devices, none had prior clinical experience using handheld spirometry or FeNO devices as part of home-based asthma diagnostic strategy; as this study examined stakeholder perspectives on emerging technologies, most survey responders would also have limited clinical experience with these technologies. Thus, the findings reflect anticipated perceptions rather than experiential insights. This study was undertaken within a broader healthcare-innovation agenda, and its insights may be transferable to future algorithm/digitally-enabled pathways. However, as the clinical utility and cost-effectiveness of home-based diagnostic testing for asthma have not yet been formally established, the findings reported here are exploratory and not intended to guide clinical practice.

## CONCLUSION

The challenges of home-based asthma diagnostics are multifaceted. A successful implementation of an effective home-based testing service must leverage multiple fundamental changes around test accessibility, resource, training and education, health disparity and patient engagement in asthma. As a critical next step, it is essential to evaluate the clinical feasibility, adherence to testing protocols followed by the estimation of test accuracies, and its clinical and cost effectiveness. To support the eventual equitable implementation, clinical studies must involve populations diverse in digital literacy, socioeconomic deprivation and educational background.

## Data Availability

Anonymised data produced in the present study are available upon reasonable request to the authors

## Acknowledgement

We would like to thank Dr Helen Ashdown (Oxford) in supporting survey distribution and steering the study progression, and Dr Tim Frank and Hannah Wardman (Manchester) in supporting the study planning and facilitating liaison with primary care. We thank our public contributors Mr Walter Kuczer and David Royle in supporting the study through patient and public involvement and engagement during the planning and study period.

