## Supplementary material for "Expectations and attitudes in primary care towards home-based testing for diagnosing asthma: a mixed methods study": Online supplemental file

**Online Supplementary materials**

Ran Wang*^1,2^, Katie Lawton*^1,3^, Binish Khatoon^1^, Joanna Smith^4^, Stephen J Fowler^1,2^, Angela Simpson^1,2^, Clare Murray^1,2^

^*^joint first authors

1. Division of Immunology, Immunity to Infection & Respiratory Medicine, School of Biological Sciences, Faculty of Biology, Medicine and Health, University of Manchester, United Kingdom.
2. Manchester University NHS Foundation Trust, United Kingdom
3. Oldham South Primary Care Network
4. School for Health and Social Care, College of Health & Wellbeing and Life Sciences, Sheffield Hallam University / Sheffield Children’s NHS Foundation Trust, Sheffield.

Corresponding author:

**Dr Ran Wang**

2^nd^ floor Education Research Centre

Wythenshawe Hospital

Southmoor road

Manchester

M23 9LT

**Section E1: Format of the focus groups**

1. The session commenced with facilitators outlining the objectives of the focus group, including the rationale for engaging healthcare professionals (HCPs) and the intended use of their feedback to inform the development of a national survey. Informed consent was obtained from all participants, including permission to audio-record the session. Subsequently, the facilitators provided a structured overview of the Nominal Group Technique (NGT) methodology to guide the discussion process.
2. A 10-minute presentation was delivered to participants summarising the National Institute for Health and Care Excellence (NICE) 2017 guidelines on asthma diagnosis (the most updated guidance at the time of the focus group activities). This was followed by a brief session led by a clinical study team member (RW), who demonstrated the diagnostic devices used in their study ([www.radica.org.uk](http://www.radica.org.uk), [Murray et al 2024]) as examples. The session included demonstrations of the fractional exhaled nitric oxide (FeNO) monitor (NOBreath®, Bedfont, Intermedical, UK) and a hand-held spirometer (Spirobank Smart®, Intermedical, UK). The facilitator departed the session upon completion of the demonstration.
3. Next, participants engaged in the silent generation of individual responses. They were asked to reflect on the prompt: “What are the potential advantages, disadvantages, barriers, and enablers for patients using home diagnostic devices for asthma?” Participants were instructed to write up to five responses for each category (advantages, disadvantages, barriers, and enablers), with each response recorded on a separate adhesive note. Emphasis was placed on eliciting independently generated responses at this stage to minimise group influence and ensure the collection of diverse individual perspectives.
4. The subsequent stage involved clarification and consolidation of the responses. Participants took turns reading aloud their first response to the prompt, proceeding sequentially until all initial responses had been shared. This process was repeated for subsequent responses until all individual contributions were displayed on a shared board. A group discussion followed, during which participants collectively reviewed the responses. Duplicate items were removed, and overlapping responses were consolidated into overarching categories through consensus. Participants then discussed and agreed upon the top ten responses for each of the four domains (advantages, disadvantages, barriers, enablers). These prioritised responses were entered into a Microsoft Forms-based ranking tool by facilitators (KL and BK). The discussions were audio-recorded to support subsequent qualitative analysis.
5. Participants were then provided with a QR code linking to the Microsoft Form, where they independently ranked the ten prioritised items for each domain according to perceived importance. Following submission, the aggregated rankings were calculated and the results were shared with the group.

**Section E2: Data analysis for free-text survey question**
As this question elicited predominantly single-word or very brief responses rather than full sentences, a simple frequency tally was conducted. All open-text answers were reviewed; responses were grouped into priority themes. There were 104 respondents to this question, yielding 105 tallies (as a small number provided more than one answer despite being asked to state a single most important factor).

***Table E1:*** National e-survey

**We are a research group based at the University of Manchester exploring whether hand-held spirometry and FeNO devices could help improve asthma diagnosis in primary care.
While asthma is the most common chronic airway disease in the UK, “one-off” clinic-based testing can result in misdiagnosis rates. Hand-held devices, with patients taking multiple readings at home could improve diagnosis rates. We have undertaken focus groups to identify the advantages, disadvantages, enablers and barriers to home testing and would like your opinion by ranking them in order of importance.
The survey will take approximately 5 minutes to complete. All potential identifiable information will be removed. If you agree to take part continue to question 1 to agree and continue to the survey.

Thank you for taking the time to complete this survey**

**Q1** Do you agree to take part in completing the survey?

- Yes
- No

Skip To: Q2 If Do you agree to take part in completing the survey? = Yes

Skip To: End of Survey If Do you agree to take part in completing the survey? = No

**Q2** Which of the following categories do you belong to?

- GP
- GP trainee
- ANP
- Nurse
- Other, please specify your role __________________________________________________

**Q3** Please provide the first 3 characters of the postcode for your practice?

________________________________________________________________

**Q4** On average how frequently do you review people that present with a likely diagnosis of asthma?

- daily
- weekly
- monthly
- yearly
- never
- Other, please specify __________________________________________________

**Q5** Please rank the barriers for using home diagnostic devices for asthma below from most important (1) to least important (10). Drag each statement to the order you would rank them.

______ Staff difficulties in interpreting results

______ Start up costs/device costs

______ Patients misunderstanding results

______ Patients without smartphones will be excluded

______ Up front training time needed for patients

______ Device might get broken/lost/stolen

______ Increased workload for staff

______ Patients may struggle with device techniques

______ Patient's compliance

______ Loss of payments for spirometry (i.e from the PCN)

**Q6** Please rank the enablers for using home diagnostic devices for asthma below from most important (1) to least important (12). Drag each statement to the order you would rank them.

______ Patients being more engaged/autonomous

______ Leaflet to explain in different languages

______ Patients that are more tech-savvy

______ Algorithm/specialist team to interpret results

______ Widely available devices

______ App reminders for patients

______ Support from hub/secondary care

______ Financial incentives

______ Training on using devices for healthcare practitioners

______ Additional staff to organise the service and work within it

______ Staff feeling they were providing better patient care

______ Training on using devices for patients for example a video/illustrations

**Q7** Please rank the advantages of using home diagnostic devices for asthma below from most important (1) to least important (12). Drag each statement to the order you would rank them.

______ More accurate diagnosis

______ Quicker diagnosis

______ Reduction in inappropriate prescribing/treatments

______ Patient-centred /personalised approach

______ Patients feeling more motivated to manage their asthma

______ Better patient health outcomes

______ Reduced referrals to secondary care

______ Reduction in use of resources i.e clinician appointments/room space

______ Capturing of diurnal variation

______ User-friendly

______ Will help diagnose children

______ Cost-effective

**Q8** Please rank the disadvantages of using home diagnostic devices for asthma below from most important (1) to least important (12). Drag each statement to the order you would rank them.

______ Cost of equipment/devices

______ Staff not feeling confident in delivering package

______ Cost of/lack of staff time to interpret results

______ Depriving patients who are not tech-savvy

______ Cost of staff training

______ Reducing other income streams

______ Having enough devices if high demand

______ Longer appointments may mean other less appointments for others

______ Increasing burden on primary care

______ Poor patient technique leading to invalid results

______ Could increase patient health inequalities

______ May not be more efficient than the status quo

**Q9** Do you think home diagnostic devices for asthma would be practical in primary care?

- Yes
- No
- Don't know
- Maybe
- Other, please specify __________________________________________________

**Q10** In your individual practice what would be the most important factor that would enable/help you to use home diagnostic devices for asthma?

________________________________________________________________

***Table E2.*** Demographic comparison of survey respondents by completion status.

|  | **Completed (included in primary analysis)**  **(n=104)** | **Not completed**  **(excluded in primary analysis)**  **(n=131)∞** | ***p*-value** |
| --- | --- | --- | --- |
| % GP, n(%) | 84/104 (80.8%) | 55/73 (75.3) | 0.745 |
| % nurses, n(%) | 9/104 (8.7%) | 6/73 (8.2%) |  |
| % ANP, n(%) | 2/104 (1.9%) | 1/73 (1.4%) |  |
| % Other, n(%) | 4/104 (3.8%) | 4/73 (5.5%) |  |
| % of practitioners who frequently care for asthma patients* | 57/104 (55%) | 40/73 (45%) | 0.269 |
| % of practitioners works in deprived area^#^ | 54/103 (52.4%) | 25/54 (46.3%) | 0.574 |
| ***** defined as more frequently than weekly encounter of asthma diagnosis in usual practice; # IMD in the lower quartile (≤3); **∞** missing data was excluded. Denominators were based on the total number of received response from the corresponding survey questions. | | | |

***Figure E1.*** Geographical locations of survey responders who did not complete the survey in full*.

***Figure E2.*** Survey respondents’ demographics of all individuals who answered the corresponding question; these were largely comparable to those who completed the survey in full.

***Figure E3.*** The ranking of the importance of potential advantages (motivators), barriers and enablers reported by e-survey of all respondents who completed the relevant question. These were largely comparable to those who completed the survey in full.
